# The timing of COVID-19 transmission

**DOI:** 10.1101/2020.09.04.20188516

**Authors:** Luca Ferretti, Alice Ledda, Chris Wymant, Lele Zhao, Virginia Ledda, Lucie Abeler-Dörner, Michelle Kendall, Anel Nurtay, Hao-Yuan Cheng, Ta-Chou Ng, Hsien-Ho Lin, Rob Hinch, Joanna Masel, A. Marm Kilpatrick, Christophe Fraser

## Abstract

The timing of SARS-CoV-2 transmission is a critical factor to understand the epidemic trajectory and the impact of isolation, contact tracing and other non-pharmaceutical interventions on the spread of COVID-19 epidemics. We examined the distribution of transmission events with respect to exposure and onset of symptoms. We show that for symptomatic individuals, the timing of transmission of SARS-CoV-2 is more strongly linked to the onset of clinical symptoms of COVID-19 than to the time since infection. We found that it was approximately centered and symmetric around the onset of symptoms, with three quarters of events occurring in the window from 2-3 days before to 2-3 days after. However, we caution against overinterpretation of the right tail of the distribution, due to its dependence on behavioural factors and interventions. We also found that the pre-symptomatic infectious period extended further back in time for individuals with longer incubation periods. This strongly suggests that information about when a case was infected should be collected where possible, in order to assess how far into the past their contacts should be traced. Overall, the fraction of transmission from strictly pre-symptomatic infections was high (41%; 95%CI 31-50%), which limits the efficacy of symptom-based interventions, and the large fraction of transmissions (35%; 95%CI 26-45%) that occur on the same day or the day after onset of symptoms underlines the critical importance of individuals distancing themselves from others as soon as they notice any symptoms, even if they are mild. Rapid or at-home testing and contextual risk information would greatly facilitate efficient early isolation.

## Introduction

The COVID-19 disease emerged at the end of 2019. Several months after the first reports on the disease, our understanding of transmission of the causative virus - SARS-CoV-2 - is still incomplete. A detailed knowledge of its transmission is urgently needed to improve public health interventions aimed at reducing the burden of the pandemic on societies. In particular, the temporal profile of infectiousness in relation to the onset of symptoms is crucial for assessing and optimising public health interventions and minimising disruption to society and the economy.

When designing interventions to control a communicable disease such as COVID-19, a key quantity is the fraction of transmissions occurring while the source is non-symptomatic (Fraser et al. 2004; Peak et al. 2017), i.e. either pre-symptomatic (before symptom onset) or asymptomatic (for individuals who never develop symptoms).

Symptomatic individuals are easier to (self-)identify and they can take measures to avoid spreading the virus, whereas transmission from non-symptomatic individuals is much more difficult to prevent. The extent and timing of non-symptomatic transmission have a large impact on which public health interventions can be effective and how they should be implemented (Fraser et al. 2004; Peak et al. 2017). For example, isolation of symptomatic cases may be sufficient to prevent spread for a disease that is transmitted only after onset of clinical symptoms, as was the case with SARS (May et al. 2004); however, further interventions may be necessary for a disease with a high proportion of non-symptomatic transmission such as COVID-19. In the latter case, preventing transmission from non-symptomatic individuals requires actively finding such individuals through contact tracing (Ferretti et al. 2020) or mass testing (Larremore et al. 2020), and/or measures targeting the entire population such as face masks, hand hygiene, and varying degrees of physical distancing. Hence, knowing the contribution of non-symptomatic transmission is key for the choice of interventions.

The fraction of all COVID-19 transmissions that come from asymptomatic individuals is difficult to measure in an unbiased manner; in addition, it is likely dependent on the age of infected individuals and therefore time- and population-specific. Indirect evidence suggests that asymptomatic individuals are less infectious than symptomatic individuals (Zhou et al. 2020; Lee et al. 2020; Madewell et al. 2020) and accounted for less than half of infections of SARS-COV-2 in China and European countries in the first half of 2020 (Pollán et al. 2020; Lavezzo et al. 2020; Buitrago-Garcia et al. 2020).

Another challenge for public health interventions is the occurrence of transmissions before or shortly after symptom onset, from individuals who do develop symptoms during the course of the infection: presymptomatic and early symptomatic transmissions hereafter. Pre-symptomatic transmissions have been estimated to account for almost half of all ever-symptomatic transmissions (see e.g. meta-analysis in (Casey et al. 2020)).

The timing of pre-symptomatic and early symptomatic transmission determines the speed required for finding the contacts who could have been infected, before these contacts, in turn, infect others. If the interval from infection to onset of symptoms is just a few days, as seems to be the case for COVID-19, it becomes critical to notify contacts instantly when the index case is confirmed by rapid testing or even symptom-based clinical diagnosis. Therefore digital contact tracing via a smartphone app, which makes the exposure notification step of contact tracing instantaneous, could substantially enhance the effectiveness of traditional manual contact tracing (Ferretti et al. 2020; Kucharski et al. 2020; Braithwaite et al. 2020; Anglemyer et al. 2020). Effectiveness of digital contact tracing depends on many factors (Hinch et al. 2020) including the fraction of the population installing the app, and on how clustered or correlated app use is within communities (Farronato et al. 2020), the time until diagnosis and notification (Kretzschmar et al. 2020), and the compliance with quarantine recommendations.

Both traditional and digital contact tracing require a reliable estimate of the temporal profile of infectiousness in order to assess which contacts are at risk of having been infected and which are not. Assessing the risk of exposed contacts as accurately as possible is key to maximising the number of infectious individuals in quarantine and preventing further spread, while minimising the number of noninfectious individuals unnecessarily quarantined.

In this study we examine the temporal profile of infectiousness for SARS-CoV-2. We estimate the distributions for the length of the intervals between infection, symptom onset and transmission using multiple datasets containing information on 191 transmission pairs. We find that the timing of transmission events depends more strongly on onset of symptoms than time since infection. Infectiousness reaches its peak near the onset of symptoms, after increasing gradually from the time of infection. We estimate the fraction of pre-symptomatic transmissions, and the relative importance of transmissions before and shortly after the time of symptom onset. Finally, we outline the relevance of these findings for contact tracing, timing of isolation and other individual precautionary measures.

## Time intervals

The temporal profile of COVID-19 infection and transmission is characterised by four epidemiologically relevant time intervals (Table 1 and Figure 1): the incubation period, the serial interval, the generation time and the time from onset of symptoms to transmission (TOST). These four time intervals are delimited by four key time points: the time at which an individual gets infected, the time at which they infect another individual, the times at which the source develops symptoms and at which the recipient does. With the exception of the incubation period, which is defined for a single individual, these intervals are defined for a transmission pair: an index (primary) case and a secondary case infected by the index.

**Table 1:** Four time intervals that influence control via isolation of symptomatic individuals.

| Time interval | From |  | To |
| --- | --- | --- | --- |
| Incubation period | infection | → | onset of symptoms |
| Generation time | infection | → | transmission (secondary) |
| TOST | onset of symptoms | → | transmission (secondary) |
| Serial interval | onset of symptoms | → | onset of symptoms (secondary) |

**Figure 1:**
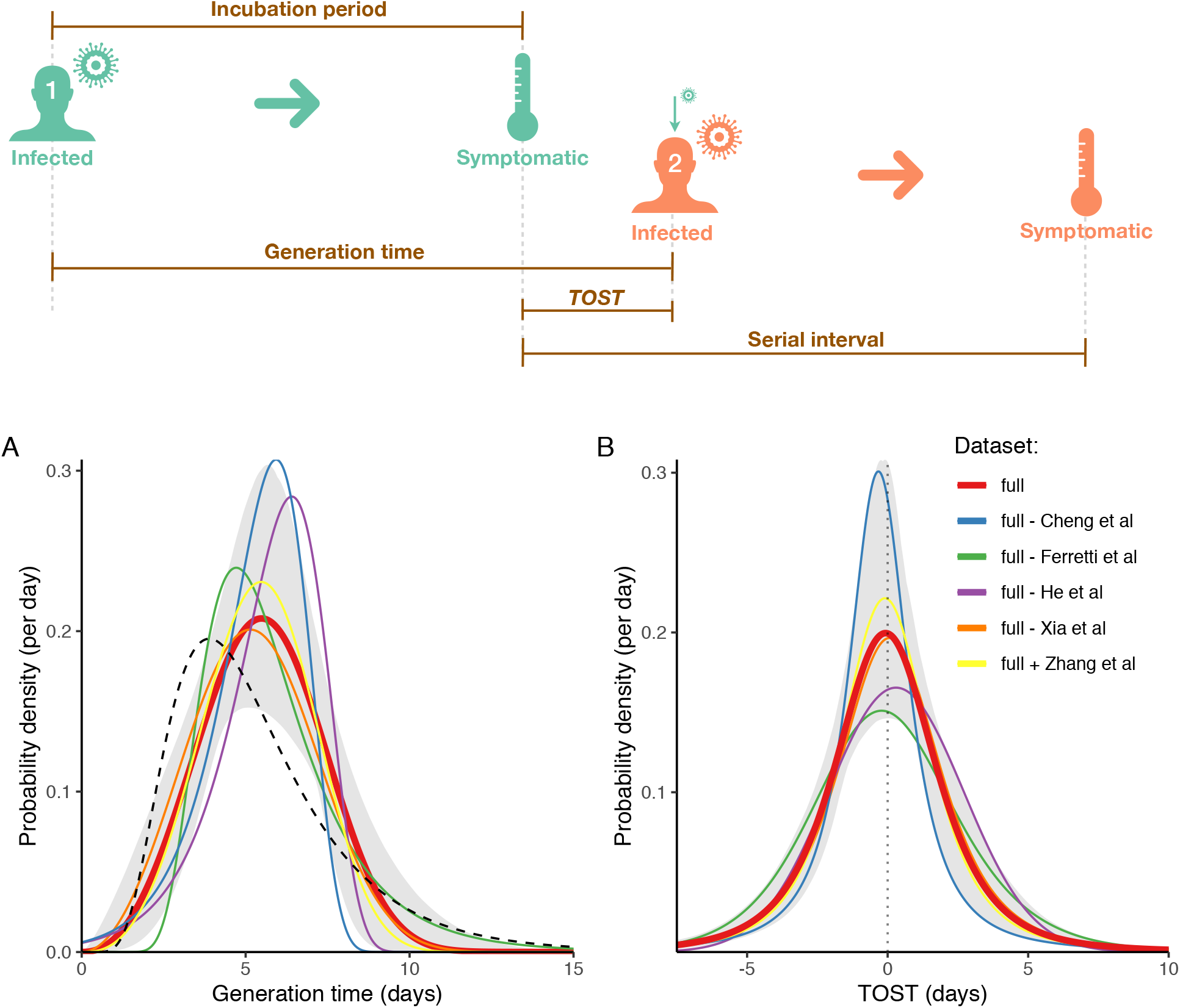
*Top: illustration of the definitions of epidemiologically relevant time intervals. Bottom: maximum-likelihood distributions of (A) generation time and (B) TOST, excluding or including different datasets, shown with different colours. The pointwise 95% CI for the fit to the baseline dataset (‘full’, which does not include Zhang et al.) is shown in grey. The dashed black line shows the incubation period distribution for comparison*.

The incubation period is the time between infection and onset of symptoms. From the average of multiple distributions from the literature (see Methods), its mean is 5.7 days and its SD is 3.5 days.

The serial interval is the interval between times of onset of symptoms in index and secondary cases. It can be directly measured from the data, and has been extensively studied for COVID-19 (Griffin et al. 2020; Nishiura, Linton, and Akhmetzhanov 2020; Du et al. 2020; Tindale et al. 2020), although it can be affected by several biases (Park et al. 2020). It is occasionally negative - when the secondary case develops symptoms before the index case. It is undefined if either case has an asymptomatic infection or does not report symptoms.

The generation time is the interval between the time of infection of the index case and the time of infection of the secondary case. It is typically harder to estimate directly unless the interval of exposure is short for both index and secondary case. The generation time is usually inferred indirectly from intervals of exposure and onset of symptoms. It has been inferred for COVID-19 by (Ferretti et al. 2020; Ganyani et al. 2020). It is always positive by definition.

TOST is the time elapsed between the onset of symptoms in the index case, and the transmission from index to secondary case. It is positive for symptomatic transmission and negative for pre-symptomatic transmission by definition. It is undefined if the index case has an asymptomatic infection or does not report symptoms. This time interval has rarely been discussed in epidemiology, although it has been considered for COVID-19 (He et al. 2020; Ashcroft et al. 2020).

Knowledge of these four intervals enables us to predict the relative effectiveness of different interventions, e.g. physical distancing, wearing of face masks, mass testing or contact tracing.

## Results

### Serial interval

We analysed transmission pairs from the only four datasets in the literature (Ferretti et al. 2020; Xia et al. 2020; Cheng et al. 2020; He et al. 2020) that contain the date of onset of symptoms for both index and secondary cases, as well as partial information on intervals of exposure. To test the robustness of the results, we included an additional dataset by (Zhang et al. 2020) with serial intervals only.

The empirical serial interval, when analysing all five datasets combined, had a mean of 5.1 days, a median of 4 days, and a standard deviation of 3.8 days; the mean was 4.1 days without Zhang et al. These results are consistent with other studies (Tindale et al. 2020; Du et al. 2020; Bi et al. 2020). Most studies fit a lognormal distribution to the data, even though serial intervals are occasionally negative. The empirical distribution and the lognormal distribution with the same mean and SD are illustrated in Supplementary Figure 1. There are, however, many caveats in using the serial interval distribution for epidemiological inference (Park et al. 2020).

The median of the serial interval was similar across different datasets (all p>0.2, two-sided Mann-Whitney U-test) but the variance was different, with Ferretti & Wymant et al and Cheng et al showing significant underdispersion and overdispersion with respect to the others (p = 0.001 and 0.025 respectively, two-sided Fligner-Killeen test). 14 out of 40 transmission events from Ferretti & Wymant et al occurred during a phase of exponential epidemic growth, when the corresponding serial interval is expected to be shorter (Svensson 2007). We explicitly corrected for this effect in the inference of generation time in the next section.

### Generation time

We inferred the generation time distribution by maximum likelihood estimation, using dates of onset of symptoms for both index and secondary cases, as well as their intervals of exposure (when available). We corrected for exponential growth and right censoring, as detailed in the Methods.

To determine the functional form of the distribution, we tested a wide range of possible shapes that are consistent with the near-absence of infectiousness of the index case at the time of infection. This requirement stems from the very low initial viral load, since each cycle of viral replication takes several hours (Bar-On et al. 2020).

The best-fitting shape, as determined by the Akaike Information Criterion (AIC), was a Weibull distribution (Figure 1A; mean 5.5 days, standard deviation 1.8 days). A Gompertz, a log-logistic and a gamma distribution provided good fits as well (*ΔAIC* = 0.57, 0.93 and 1.06 respectively, Supplementary Table 1). The generation time distributions corresponding to the best parameter fits for each shape are shown in Supplementary Figure 2, and the maximum-likelihood parameter values for each shape in Supplementary Table 2.

Given the heterogeneity among datasets in terms of interventions and stage of the epidemic, it is important to assess the robustness of the inference with respect to the choice of datasets. We re-inferred the generation time distribution removing one dataset at a time, as well as adding the additional dataset by Zhang et al. The results were quite robust and within the uncertainties of the inference, as illustrated in Figure 1A.

### Time from onset of symptoms to transmission (TOST)

We inferred the distribution for TOST by maximum likelihood estimation. As COVID-19 is often transmitted pre-symptomatically (negative TOST), we considered distributions which include both positive and negative times. The best fit was given by a scaled Student’s *t* distribution (Fig. 1B; mean = −0.07 days, SD = 2.8 days). A skew-logistic distribution (mean = 0.02 days, SD = 2.65 days) also provided a good fit (*ΔAIC* = 0.65; Supplementary Table 3). The TOST distributions corresponding to the best parameter fits for each shape are shown in Supplementary Figure 3, and the parameter values in Supplementary Table 4.

We tested the robustness with respect to choice of datasets by removing one dataset at a time and adding an additional dataset as before. All best-fit distributions were centered around the time of onset of symptoms with roughly similar width (Figure 1B).

#### The peak of infectiousness depends on the onset of symptoms rather than the time of infection

It has previously been noted that the similarity between the mean incubation period and the mean generation time (about 5.5 days) suggests a mean TOST of close to zero i.e. an almost-centered distribution of TOST (Nishiura, Linton, and Akhmetzhanov 2020; Casey et al. 2020). Transmission events are indeed clearly concentrated around the time of onset of symptoms, regardless of the details of the inference. This can be either a coincidence, with no relation between generation time and incubation period (symptom-independent infectiousness), or the time since onset of symptoms could determine the infectiousness of an individual, rather than the time since the individual was first infected (symptom-dependent infectiousness). These two scenarios are illustrated in Figure 2A and 2B respectively.

**Figure 2:**
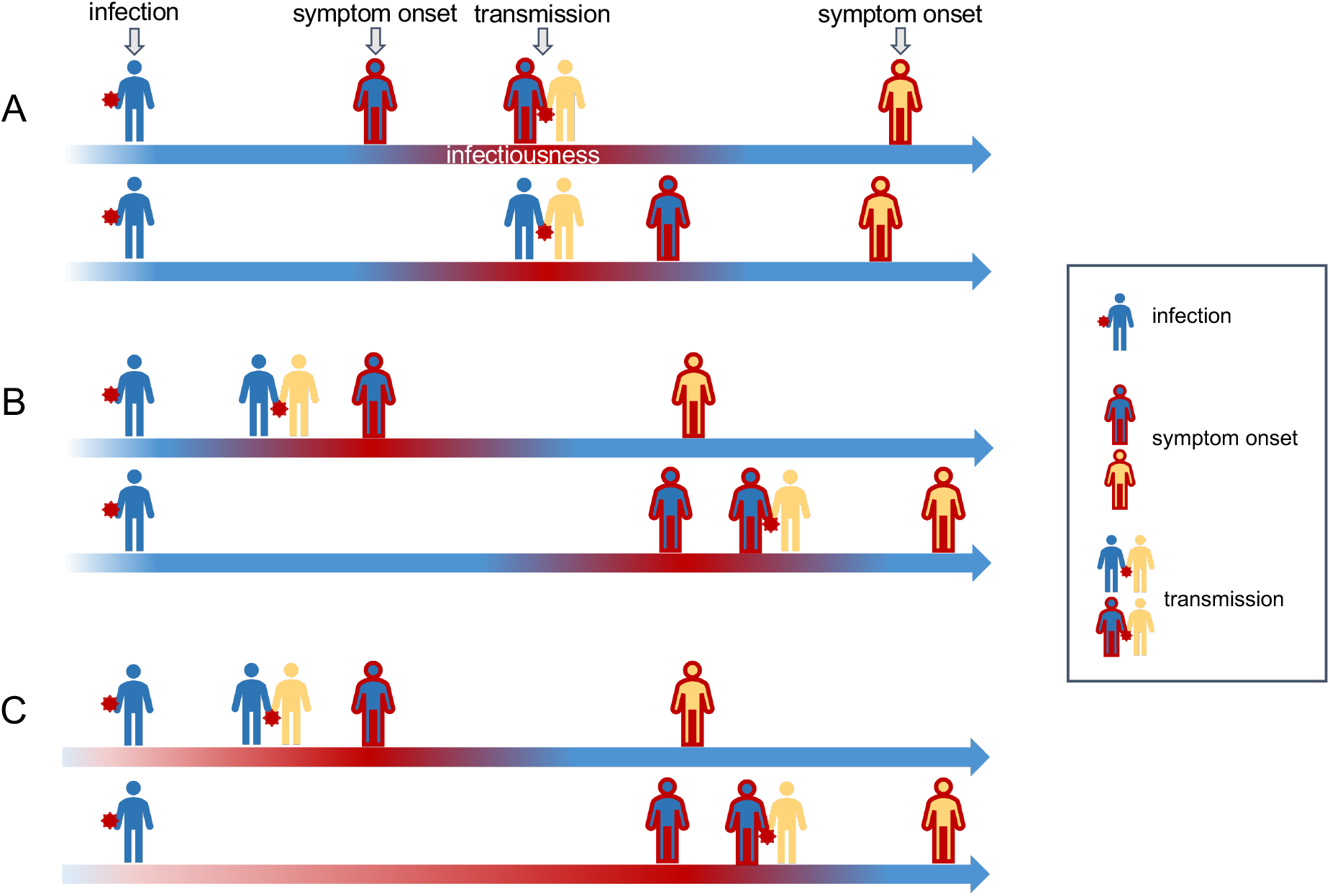
*Three alternative hypotheses for the timing of infectiousness of the blue individual (and individuals generally). In (A), infectiousness depends on the time since infection, regardless of whether symptoms develop quickly (A top) or slowly (A bottom). In (B), infectiousness depends on the time of the onset of symptoms, regardless of whether the time since infection is short (B top) or long (B bottom). In (C), infectiousness depends on the time of onset of symptoms, but it increases gradually from the time of infection, therefore leading to a shorter infectious period if symptoms develop rapidly (C top) and a longer infectious period if symptoms appear late (C bottom)*.

We tested which of these two scenarios, i.e. symptom-dependent and symptom-independent infectiousness, best fit our data on transmission pairs, using the information on the interval of exposure of the index and secondary cases compared with the time of onset of symptoms of the index. Comparing the AIC values in Supplementary Tables F and G for the most directly informative datasets (Ferretti & Wymant et al, Xia et al, and both combined), we found clear differences between the best fits for symptom-dependent and symptom-independent infectiousness (*ΔAIC* = −13.9, −11.1, −22.4 for the different datasets respectively; Supplementary Table 5). Even the simplest Gaussian fit for symptom-dependent infectiousness was a much better fit to the data than any choice of generation time distribution for symptom-independent infectiousness (*ΔAIC* = −5.4, −11.1, −18.8).

These results confirm the observation that for individuals that eventually develop symptoms, the period of SARS-CoV-2 infectiousness is directly related to onset of symptoms rather than being independent of it. For symptomatic individuals, most transmission events occurred in a range of a few days before and after onset of symptoms. More than 5 days before symptom onset, infectiousness appeared to decrease below a tenth of its peak value, and we observed only a few percent of transmissions beyond 5 days after symptom onset.

### Infectiousness increases gradually from time of infection to onset of symptoms

Our analyses suggest that time of symptom onset is the main determinant of when transmission occurs, with transmission peaking before and after the onset of symptoms. However, the incubation period could still affect the width of the distribution of the time of transmission events. Indeed, the data suggest a weak negative correlation between the incubation period of the source and the serial interval of the pair (Supplementary Figure 4), which would not be expected if the timing of transmission was determined only by the TOST (see Methods).

To examine the possibility that the length of the infectious period depends on the incubation period (incubation/symptom-dependent infectiousness), we considered several possible dependencies between the TOST distribution and the incubation period. We modelled them by rescaling the time of transmission by a factor dependent on the incubation period, affecting either the whole distribution or its left part only (details in Methods and Supplementary Table 5).

The model of incubation/symptom-dependent infectiousness with the best fit across all datasets was a linear rescaling of pre-symptomatic values of TOST by the length of the incubation period (Figure 2C; *ΔAIC* = 0, −9.3, −10.2, −15.5 for Ferretti & Wymant et al, Xia et al, both combined, and all datasets combined, respectively). In other words, individuals with longer incubation periods also tend to have a proportionally earlier and longer pre-symptomatic infectious period (Figure 3A). The profiles of transmission for different incubation periods in relation to TOST are depicted in Figure 3B. The best fit for TOST is provided by a rescaled skew-logistic distribution as a function of the TOST *t* and the incubation period *t*_*i*_:

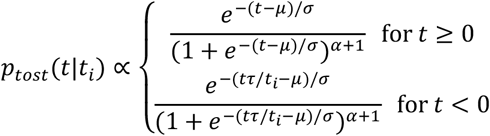

with parameters *μ* = −4.00 days, *σ* = 1.85 days, *α* = 5.85. The constant *τ* = 5.42 days is the mean incubation period according to (McAloon et al. 2020). For an incubation period *t* = *τ*, the TOST has a mean of 0.1 days and SD of 2.4 days.

**Figure 3:**
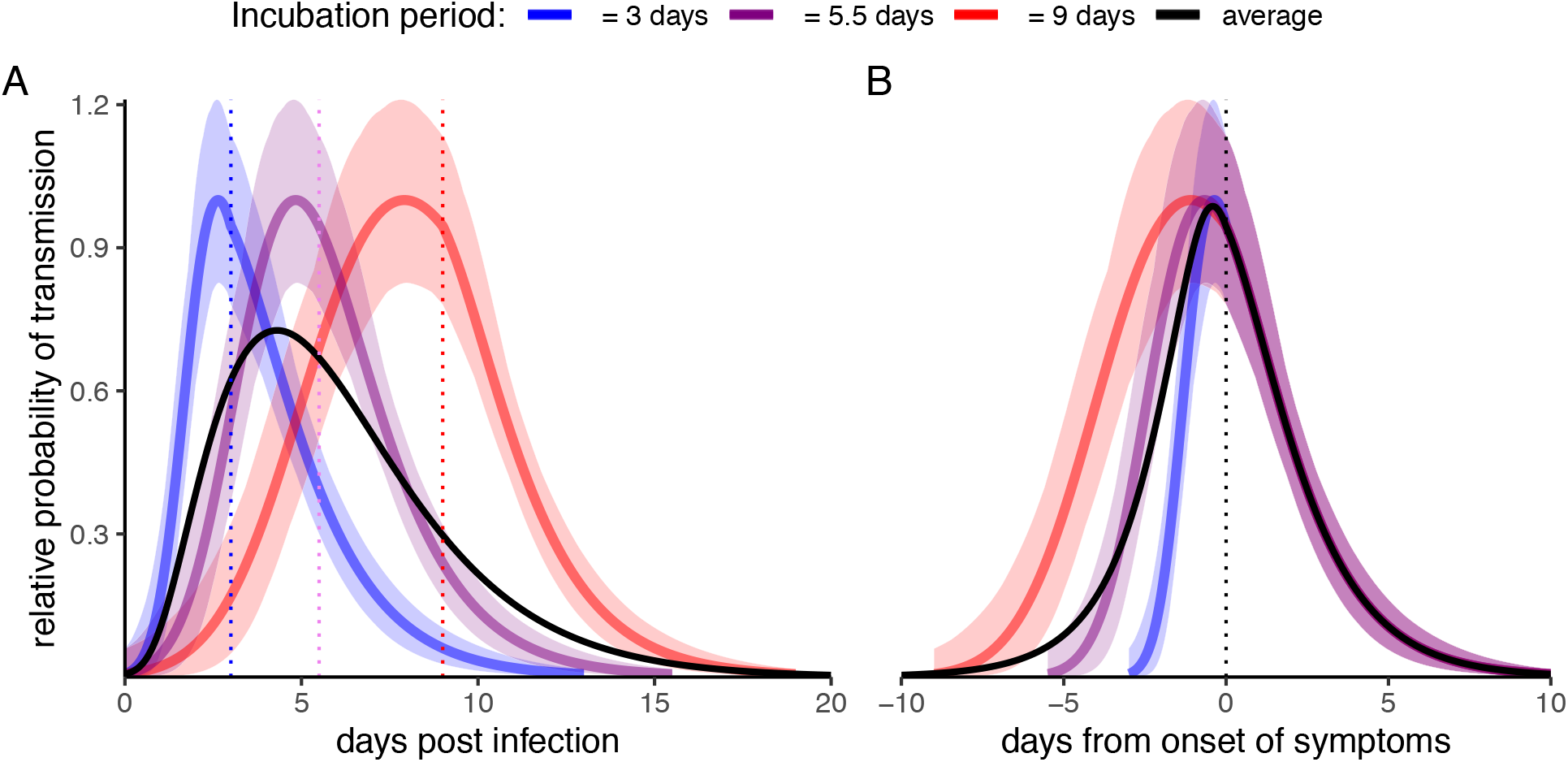
*probability of transmission as a function of generation time (left) and TOST (right) for a given duration of the incubation period, relative to the peak probability. The black line represents the average with respect to the incubation period distribution. The envelopes correspond to the pointwise 95% CI*.

It is unclear whether the magnitude of infectiousness depends on the incubation period. It is possible that an early host immune response could initially reduce viral replication, but once initial barriers of immunity are overcome, the same peak infectiousness could be reached. This would mean that individuals with later onset of symptoms have a longer infectious period with the same peak infectiousness; i.e. their cumulative infectiousness increases with the duration of their incubation period. Our dataset cannot discriminate between this scenario and the one where cumulative infectiousness does not change depending on the length of the incubation period (Supplementary Table 5). Irrespective of this, the average distribution of TOST over all durations of the incubation period is very similar to the previous fit (Supplementary Figure 5).

Since the time of symptom onset is the main determinant of infectiousness, we notice a strong positive correlation between incubation period and generation time, which is clearly visible in Figure 3A, and a weaker negative one between incubation period and TOST (Supplementary Table 6).

### Epidemiological biases could affect symptomatic transmissions

Many biases can potentially affect the shape of the temporal profile of transmissions inferred from observed transmission pairs These include both sampling biases and self-isolation or public health interventions.

The more time has passed since an individual became infected, the more likely they are to have developed symptoms or have a positive test result (either through mass testing or as a traced contact), and thus the more likely they are to have begun taking precautions to avoid transmitting the virus. Hence, inferred distributions of transmission events could be prematurely truncated relative to the distribution that would be observed with no interventions, and therefore differ from the profile of purely biological infectiousness over time.

Such epidemiological biases also affect the right tail of the distribution of serial intervals. In particular, the serial interval can change due to interventions (Ali et al. 2020)(Sun et al, personal communication) as it depends strongly on time to isolation (Bi et al. 2020). These biases cannot be disentangled using data from transmission pairs only, but require information on dates of exposure for traced contacts who were exposed but did not get infected. We extended our approach to include such information from (Cheng et al. 2020) (Supplementary Figure 6), fitting a model with dataset-dependent decay of transmissions after symptoms (Supplementary Methods). The resulting distribution of TOST does not differ from our previous fit, lying mostly within its confidence intervals (Supplementary Figure 7). Furthermore, the right tail of symptomatic transmissions is identical for all datasets. These results confirm that the distribution of TOST is robust and that epidemiological biases are similar between the studies, such as the tendency to self-isolate when experiencing acute respiratory symptoms.

We caution, however, that infected individuals could be still infectious well beyond the time periods suggested by this epidemiological analysis. Viral loads from nasal swabs provide substantial evidence of an infectious period after symptom onset longer than the 3-4 days inferred here. High viral loads have been observed for at least a week after symptoms (Wölfel et al. 2020; Pan et al. 2020; He et al. 2020). The crucial determinant of infectiousness however is the probability of shedding *viable* virus, which decays rapidly after 5-10 days (Wölfel et al. 2020; Bullard et al. 2020). However, Supplementary Figure 7 clearly shows that both viral load and the probability of viable virus isolation from multiple studies (Arons et al. 2020; van Kampen et al. 2020) decay more slowly than our epidemiological observations suggest. Resolution of this mismatch is likely accounted for by the increased compliance to isolation after onset of symptoms, discussed above.

### Fractions of pre-symptomatic and early symptomatic transmissions

A key feature of COVID-19 spread is pre-symptomatic transmission. Several studies have estimated its fraction by assuming time to symptoms and time to transmission are independent (Ferretti et al. 2020; Casey et al. 2020). As we have shown, this assumption is not supported by data: transmission events are closely tied to symptom onset. Also, transmission on the day of symptom onset is not necessarily presymptomatic and should be considered separately. Here we assess the contribution of strictly presymptomatic transmission using two different approaches.

First, we estimated the fraction of transmission on each individual day by discretizing the TOST distribution and considering the part of the distribution that corresponds to negative times, i.e. the fraction of transmission events that occur before the day of symptom onset (Figure 4).

**Figure 4:**
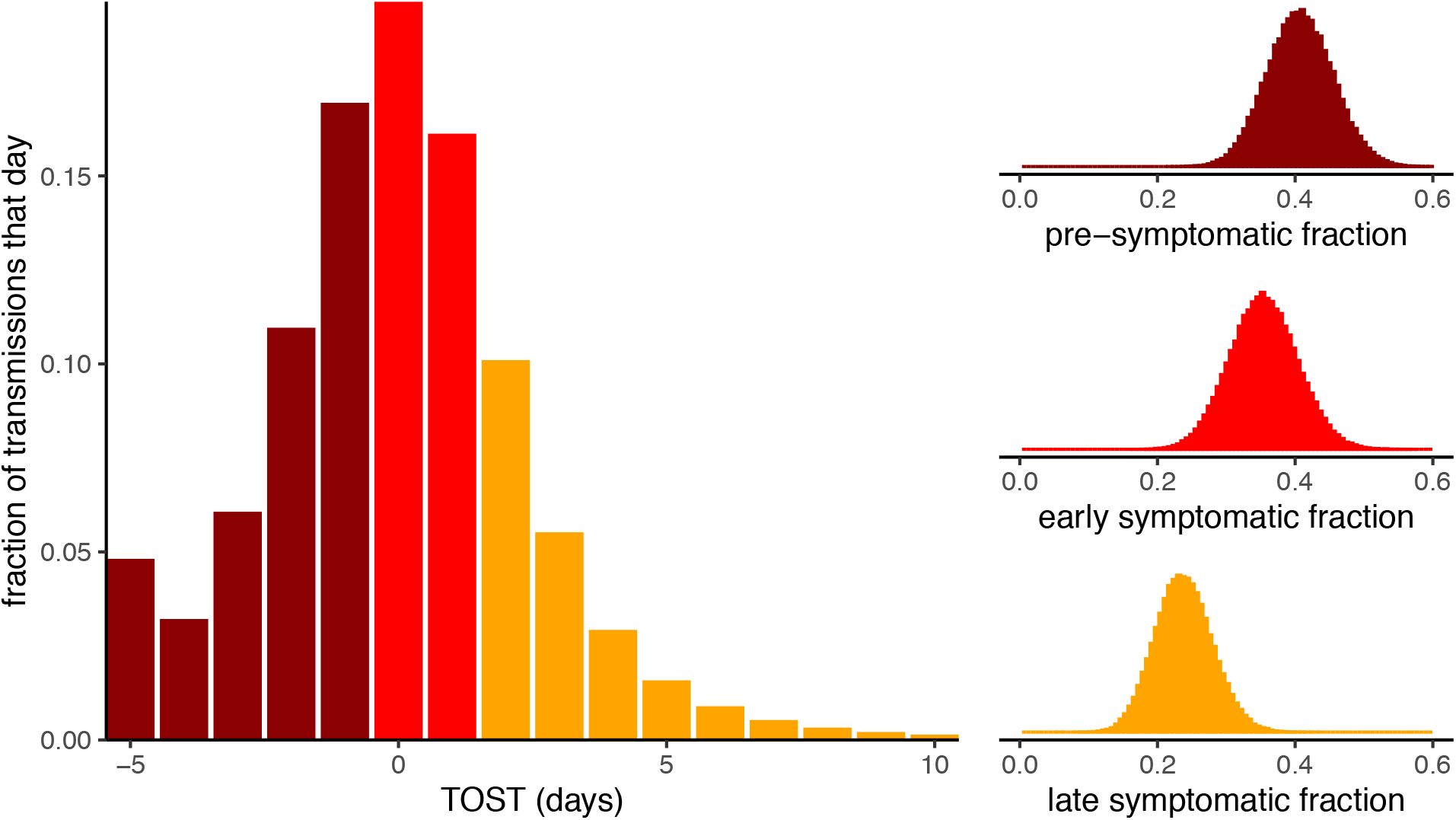
*Left: distribution of transmissions relative to the day of onset of symptoms. The left-most bin contains all transmission 5+ days before symptom onset. Right: posterior distributions of the fraction of all transmissions that occur before symptoms (pre-symptomatic, TOST<0), on the day of onset of symptoms or the following day (early symptomatic, TOST = 0-1) or thereafter (late symptomatic, TOST>1), obtained from 10000 bootstraps from all pairs in the full dataset*.

The fraction of strictly pre-symptomatic transmissions (TOST<0) from ever-symptomatic individuals was 42%, which is consistent with previous studies, even if some of them included the same day as symptom onset (TOST = 0).

We also estimate the fraction of pre-symptomatic transmissions among all pairs in our dataset with a Bayesian approach, assuming that pre-symptomatic infectiousness would scale with the incubation period (i.e. our best model, illustrated in Figure 3). This approach also estimates the fraction of presymptomatic transmissions to be 41% (95% CI: 31-50%; the full distribution is shown in Figure 4).

While much attention has been focused on pre-symptomatic transmission, the contribution of early symptomatic transmission is also crucial for the spread of the disease.

The peak of transmission occurs on the day of symptom onset, with an estimated 20% of transmissions, as illustrated in Figure 4. The day after onset of symptoms is also very relevant, accounting for 16% of transmissions. Together, these two days account for about a third of ever-symptomatic transmissions, comparable to the fraction of pre-symptomatic transmission. This estimate is confirmed by the Bayesian analysis of individual pairs in our datasets which gives a value of 35% (95% CI: 26-45%).

In contrast, symptomatic transmissions occurring two days or more after onset of symptoms account for only 22% of transmissions (24% from Bayesian analysis, 95% CI: 16-32%), although this value is likely to be affected by self-isolation and non-pharmaceutical interventions as discussed before.

The above results demonstrate the importance of implementing non-pharmaceutical interventions to reduce pre-symptomatic transmission, such as mass testing, contact tracing and physical distancing.

They also underline the importance of strict infection control measures at the first sign of even mild symptoms potentially related to COVID-19 (such as cough, fever, fatigue or anosmia), in order to reduce early symptomatic transmission. According to our results, perfect isolation of cases from onset of symptoms would stop twice as many transmissions compared to isolation from the second day after onset of symptoms, relative to a baseline with no intervention at all (excluding transmissions from fully asymptomatic individuals). Self-isolation of symptomatic individuals is therefore especially important for the first two days.

Instant, universal, and perfect self-isolation, including from family members, is challenging, given the low specificity of early COVID-19 symptoms and the high prevalence of respiratory viruses with similar symptoms between autumn and spring (Menni et al. 2020). Nevertheless, if low-cost good practices that are widely advisable irrespective of symptoms − wearing a face mask, increasing spatial distance, practicing enhanced hygiene (especially hand hygiene), and limiting social contacts (including staying away as much as possible from offices, schools, public transport, and closed public spaces) − were followed strictly at the first onset of symptoms, even if mild, this could have a substantial impact on the epidemic. Such a policy would greatly depend on compliance and collaboration from the public. Symptom tracker apps (Drew et al. 2020) could play a role in enhancing public awareness of mild COVID-19 symptoms and compliance. As a further advantage, this policy could also reduce the burden of other respiratory viruses.

Reducing barriers to testing, and increasing the speed of results is also critical; mass testing with home-based or point-of-care rapid testing could help individuals respond appropriately and quickly to onset of mild symptoms (Larremore et al. 2020). Likewise, exposure notification by contact tracing, and any information that can be provided that will help individuals gauge their local risk, will help individuals correctly interpret the onset of initial symptoms that might be very non-specific at initial onset.

## Discussion

We have presented an in-depth analysis of the timing of transmission from a selection of the most informative datasets of transmission pairs currently available. The resulting picture of the temporal infectiousness profile of COVID-19 has some clear consequences in terms of policy.

The most immediate consequences concern the assessment of the transmission risk associated to contacts for both manual and digital tracing or exposure notification approaches. Contact tracing guidelines from the CDC (CDC 2020) and the WHO (WHO 2020) support tracing of contacts up to two days before the onset of symptoms or, for asymptomatic individuals, before a positive test. For symptomatic individuals, this approach misses approximately 10% of all transmissions overall, but a much larger fraction for individuals with a longer incubation period (Supplementary Figure 8). Hence, public health advice should consider appropriately longer intervals for contact tracing whenever an estimate of the date of infection is available (e.g. through backward contact tracing) and it precedes symptoms by more than 4-5 days.

For digital contact tracing, which relies on algorithms for risk scoring that can take into account the time profile of infectiousness (Wilson et al. 2020), our results are highly valuable to inform the appropriate risk scoring algorithm. They show that the date of onset of symptoms for the index case is a critical piece of information for the algorithm, which should be collected preferably within the app itself, or by public health officials at the time the case is confirmed.

Before symptom onset, relative transmission probabilities estimated here can be used as a proxy for infectiousness, i.e. the degree of danger from a given exposure. After symptom onset, risk scores should take into account that the distribution inferred here is likely to underestimate infectiousness. We note that quantitative treatment of infectiousness is only possible in early versions of the Google/Apple exposure notification system; unfortunately, recent versions do not permit more than two levels of infectiousness.

The profile of infectiousness possibly depends also on other parameters, such as infection severity and age. The infectiousness profile for fully asymptomatic individuals is unknown. For symptomatic individuals, the limited data available do not suggest a strong dependence on age (Ali et al. 2020; Furuse et al. 2020), but further studies are needed.

The definition of the date of onset of symptoms suffers from uncertainties related to ambiguities in the choice of the set of symptoms associated to COVID-19, the time of onset of different symptoms, and the uncertainties in their recall. All these sources of uncertainty are implicitly present in our study as well. Given a known, fixed, and identifiable set of symptoms, the date of onset of symptoms can be retrieved with reasonable precision. However, the lack of specificity for COVID-19 symptoms, international variation in recognised symptoms, and reliance on patients’ recollections all present challenges. The date of infection often has an even greater degree of uncertainty due to the challenges in identifying infectors (who could be non-symptomatic) and in assessing the number and duration of exposures. When the profile of infectiousness as a function of TOST is used for risk scoring, uncertainties in the date of symptom onset should be taken into account (e.g. averaging the index’s expected infectiousness at the time of exposure over the window of possible times of symptom onset).

The large fraction of transmissions that occur either before or shortly after onset of symptoms confirms that isolation of cases more than 2 days after onset of symptoms is insufficient to control the epidemic. Physical distancing, mask wearing, community testing and contact tracing are key non-pharmaceutical interventions that are able to stop transmissions before and around the onset of symptoms, and should be included in any effective strategy against COVID-19.

Current physical distancing policies in many countries already include self-isolation after COVID-19 symptoms, reduction of social interactions, and use of face masks in public places. Our results on early symptomatic transmission underline the importance of following existing guidelines as strictly as possible for the first two days of symptoms, even if symptoms are mild or it is unclear whether they are compatible with COVID-19. A policy based on reinforcing official advice or suggesting to take extra precautions for the first two days of symptoms is advisable and likely beneficial with minor economic and social costs.

## Materials and Methods

### Relation between generation time, incubation period and TOST

The generation time *t*_*g*_, incubation period *t*_*i*_ and time from onset of symptoms to transmission (TOST) *t*_*ost*_ by definition satisfy the equation

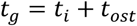

In terms of Pearson’s correlation between these epidemiological variables, this implies

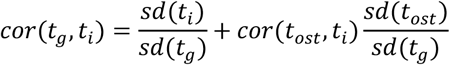

If the generation time is independent of the incubation period, this would imply an anticorrelation between the latter and the TOST, i.e.

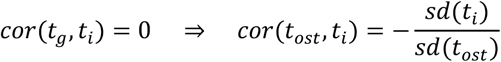

On the other hand, if the TOST is independent of the incubation period, this implies a positive correlation between generation time and incubation period

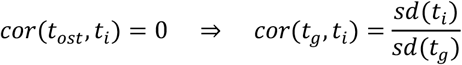

Therefore, if index cases’ incubation periods correlate more strongly with their generation times, this suggests infectiousness is driven more strongly by TOST; if they correlate more strongly with TOST, this suggests that infectiousness is driven more strongly by time since infection. Equivalently, if a model with independent distributions for TOST and incubation period better describes the pattern of infectiousness in the data than one with independent generation time and incubation period, this strongly suggests a correlation between the generation time and the incubation period of the disease. Assuming no correlation in the incubation periods of source and recipient (no heritability), it would also imply a null correlation between the incubation period of the source and the serial interval:

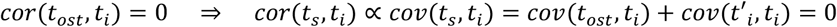

We define the distributions for *t*_*g*_, *t*_*i*_ and *t*_*ost*_ to be *p*_*g*_, *p*_*i*_ and *p*_*ost*_ respectively. It is well known that the generation time distribution *p*_*g*_(*t*) plays a key role in the renewal equation for incidence *I*(*t*) = 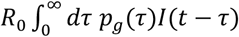 (Wallinga and Lipsitch 2007; Fraser 2007). Similarly, the TOST distribution *p*_*ost*_(*t*|*t*_*i*_) for a specific incubation period *t*_*i*_ appears naturally in an alternative renewal equation relating the number of newly symptomatic individuals at time *t* with incubation period *t*_*i*_, *C*(*t*,*t*_*i*_) to the number of new infections at time *t* from infectors with incubation period *t*_*i*_, *I*(*t*,*t*_*i*_):

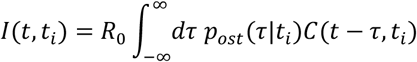

The ansatz of *I*(*t*, *t*_*i*_) exponentially growing with *t* leads to an alternative version of the Euler-Lotka equation relating *R*_*0*_ and individual transmission timing to the exponential growth rate *r*:

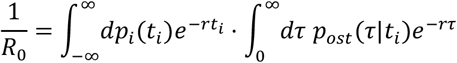

This equation neglects asymptomatic infections.

### Data and datasets

Estimations of generation time often use information on the dates of onset of symptoms for both index and secondary cases from transmission pairs (i.e. the empirical serial intervals), combined with knowledge of the incubation period distribution (Ferretti et al. 2020; Ganyani et al. 2020). However, an implicit assumption in this approach is independence between incubation period and generation time. Since this is precisely the assumption being tested in this work, we restricted our analysis to more informative datasets. To avoid issues with potential misassignment of the index case, we discarded transmission pairs from (He et al. 2020; Xia et al. 2020) belonging to clusters with more than one secondary case.

For a direct assessment of the relationship between *t*_*i*_, *t*_*g*_ and *t*_*ost*_, information on timing of exposure for both index and secondary case is needed, as well as date of symptom onset for the index case. We were able to find only two datasets that provide both the dates of onset of symptoms and (partial) information on exposure intervals for both the index case and the secondary case, namely (Ferretti et al. 2020) (40 pairs from different geographic areas), and (Xia et al. 2020) (32 pairs from China).

To further improve the accuracy of the inferred distribution of TOST, we also included datasets providing dates of onset of symptoms for the index cases as well as intervals of exposure for the secondary cases, namely (He et al. 2020) (66 pairs from China) and (Cheng et al. 2020) (18 pairs from Taiwan, excluding asymptomatic secondary cases). The latter also provides dates of onset of symptoms and intervals of exposure for contacts that did not lead to infections (2740 pairs).

As a check of robustness of our selection of datasets, we also included data on serial intervals from (Zhang et al. 2020) (35 pairs from China). There could be some overlap among the three datasets of Chinese pairs; however, the empirical serial interval distributions are sufficiently different to assume that any overlap represents a minority of transmission pairs.

For the incubation period, we considered a meta-distribution obtained by averaging seven lognormal distributions reported in the literature (Bi et al. 2020; Lauer et al. 2020; Li et al. 2020; Linton et al. 2020; Ma et al. 2020; Zhang et al. 2020; Jiang et al. 2020). The probability of onset of symptoms *P*_*i*_(*t*) at day *t* was obtained by integrating this distribution between *t*−0.5 and *t*+0.5. To test the robustness of the results, we replicated them using a lognormal distribution with mean 5.42 days and standard deviation 2.7 days, following the meta-analysis (McAloon et al. 2020). The latter provided a worse fit to our data (Supplementary Table 5), but confirmed all our conclusions; in practice, the difference between the inferred distributions was small (Supplementary Figure 5).

### Likelihood

We used a maximum composite likelihood approach similar to previous work on COVID-19 (Ferretti et al. 2020; He et al. 2020). Assuming that the transmission probability per contact per day is small, the likelihood function for a transmission pair is

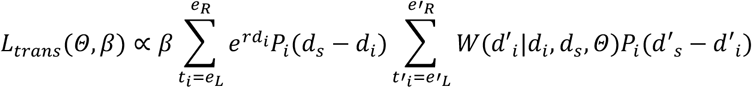

where *e*_L_, *e*_R_ are the extremes of the interval of exposure for the index case, *d*_*i*_ and *d*_*s*_ are the dates of infection and onset of symptoms for the index case, and *e*′_L_, *e*′_R_, *d*′_*i*_, *d*′_*s*_ are the same quantities for the secondary case. The parameters *β* (absolute infectiousness) and Θ (set of parameters of the time distribution) depend on the dataset. We considered multiple choices for the discretised infectiousness profile *W*(*d*′_*i*_|*d*_*i*_, *d*, *Θ*) = *Ω*_*gt*_(*d*′_*i*_ − *d*_*i*_|*Θ*) and *W*(*d*′_*i*_|*d*, *d*_*s*_, *Θ*) = *Ω*_*TOST*_(*d*′_*i*_ − *d*_*s*_|*Θ*). The growth rate *r* of the epidemic was taken to be 0.14/day for those transmission pairs sampled from the early Chinese outbreak as explained in (Ferretti et al. 2020), and 0 otherwise. The factor *e*^*rd*_*i*_^ weights an otherwise uniform distribution for *d*_*i*_ within the exposure window [*e*_*L*_, *e*_*R*_], to correct for the fact that growing incidence in the wider population makes more recent infection more likely (except when conditioning on a known infector, as for the secondary case).

Finally, the likelihood is the product of the individual likelihoods over all transmission pairs. For the inclusion of case-contact pairs from (Cheng et al. 2020) where transmission did not occur, each such pair contributes a multiplicative factor 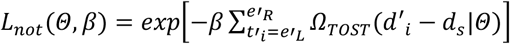 to the likelihood.

### Choice of distributions

For the generation time, we tested lognormal, gamma and Weibull distributions previously used in other studies, as well as generalised gamma, Gompertz, inverse gamma, log-logistic, Frechet, Beta’, and Levy distributions. Discretised distributions as a function of the number of days *d* were obtained by integrating each distribution between *d−0.5* and *d+0.5*.

For TOST, we considered the normal distribution, distributions with more weight in the tails (rescaled Student’s t, rescaled Cauchy), and asymmetric distributions with Gaussian and exponential tails (skew-normal, skew-logistic). We considered both the naive distribution and a version truncated at the time of infection (Supplementary Table 5); the difference between inferred shapes was small (Supplementary Figure 5).

For the analysis of the TOST distribution with different tails, we considered a range of shifted and rescaled symmetric functions (normal, generalised normal, Student’s t, Cauchy) and we modelled the left and right side of the peak separately, assuming a continuous probability density.

Finally, we modelled several options for joint distributions by rescaling the values of TOST depending on the incubation period. More specifically, we either rescaled the TOST either relative to the onset of symptoms or to the location parameter of the distribution, and we rescaled either both tails or the left tail only. We also performed some more complex rescalings (Supplementary Table 5 and Supplementary Methods). Note that when conditioning on the incubation period, the left tail of TOST distributions was truncated at the time of infection.

### Bayesian reconstruction of timing of transmission for individual pairs

For each transmission pair, we assumed an equal prior probability of transmission on any given day. The pair’s posterior probability of pre-symptomatic transmission therefore is the likelihood of transmission conditional on being pre-symptomatic, divided by the overall likelihood of transmission. To estimate the uncertainty on the overall fraction of pre-symptomatic transmissions, first we resampled the same number of pairs at random with replacement, then we assigned each pair as pre-symptomatic transmission or not according to the abovementioned posterior; we repeated the process 10,000 times to obtain the corresponding empirical distribution. We proceeded in the same way for early and late symptomatic transmissions.

## Data Availability

All datasets used in this paper are published or publicly available.

## Acknowledgments

We thank Katrina Lythgoe and the Oxford Pathogen Dynamics Group for useful discussions and Claudio Ferretti for help with data extraction. The study was funded by an award from the Li Ka Shing Foundation to CF. The funders of the study had no role in study design, data collection, data analysis, data interpretation, or writing of the report.

## Author Contributions

Conceptualization: LF, CW, JM, AMK, CF. Data curation: LF, VL, HC, TN, HL. Funding acquisition: CF. Investigation: LF, AL, CW. Methodology: LF, AL, CW, RH. Visualization: LF, AL, CW, LZ, MK. Writing, original draft: LF, AL, CW, LAB, AN, JM, AMK, CF. Writing, review, and editing: all authors.

## Data availability

Data is available from the original publications (Ferretti et al. 2020; Xia et al. 2020; He et al. 2020; Cheng et al. 2020; Zhang et al. 2020; Arons et al. 2020; Van Kampen et al. 2020). Processed data is also available at 10.5281/zenodo.4033022.

## Code availability

The R code used for the analysis is available at 10.5281/zenodo.4033022.

